# Optimal Thresholds for Cardiac CT Parameters in Patients with Aortic Stenosis Referred for Transcatheter Intervention

**DOI:** 10.64898/2026.01.07.26343641

**Authors:** Tarun K Mittal, Kivraj Sabarwal, Ben Ariff, Saeed Mirsadraee, Sandeep S Hothi

**Author notes:** Corresponding author: Tarun K Mittal MD, FRCR Department of Imaging & Cardiology, Harefield Hospital, Guy’s & St. Thomas’ NHS Foundation Trust, Hill End Road, Uxbridge UB9 6JH London, UK.

## Abstract

**Background:** Computed tomography (CT) is routinely performed for planning transcatheter aortic valve implantation (TAVI) procedures. Aortic valve calcium score (AVCS) is recommended to ascertain the severity of degenerative aortic stenosis (AS) in low-flow states, with the role of CT-AVA (aortic valve area) being uncertain. In this study, we determined the best AVCS and CT-AVA thresholds in patients referred for TAVI.

**Methods:** A retrospective multicentre study evaluated 1162 patients undergoing TAVI CT for severe AS. Two inclusion criteria for normal cardiac output by transthoracic echocardiography (TTE) were considered: Group A: left ventricular ejection fraction ≥50% and indexed stroke volume 28-48 ml/m^2^, Group B: cardiac index 1.9-4.3 L/min/m^2^. The predictive ability of CT parameters for severe AS (mean gradient ≥40 mmHg) was assessed using ROC curves and optimal thresholds were determined.

**Results:** 428 patients in Group A and 685 patients in Group B fulfilled the inclusion criteria. Best thresholds for AVCS and CT-AVA in group A (mean age 81±7.5 years; 54% women) were ≥2034 AU (AUC 0.79) and ≤0.90 cm^2^ (AUC 0.71) for women and ≥3046 AU (AUC 0.78) and ≤0.97 cm^2^ (AUC 0.69) for men; while in Group B (mean age 81±7.5 years; 45% women), they were ≥1962 AU (AUC 0.80) and ≤0.99 cm^2^ (AUC 0.73) in women, and ≥2983 AU (AUC 0.78) and ≤1.04 cm^2^ in men.

**Conclusions:** This study defines optimal AVCS thresholds for severe AS in a TAVI patient cohort, which are much higher than previously reported. CT-AVA could also be an additional parameter to establish AS severity.

## INTRODUCTION

Aortic stenosis (AS) is the commonest primary adult heart valve disease in Western countries with increasing prevalence in those aged over 65 years.^1,2^ The diagnosis of severe AS is made with transthoracic echocardiography (TTE) as the primary imaging modality, using the peak velocity (Vmax) of blood flow through the valve, mean pressure gradient (MG), or the effective valve area (EVA) calculated using the continuity equation.^3,4^ Once the diagnosis of severe aortic valve (AV) stenosis is confirmed in a symptomatic patient or in an asymptomatic patient with left ventricular systolic dysfunction without another cause, treatment is with surgical aortic valve replacement (AVR) or, increasingly, with trans-catheter aortic valve implantation (TAVI) in patients at intermediate to high risk for surgery or, in recent ESC/EACTS guidance, TAVI is now favoured in patients ≥ 70 years.^5,6^

However, Vmax and MG are both flow-dependent parameters that can underestimate the degree of AS in low flow states.^3–7^ and can overestimate severe AS in high flow states.^8^ In patients with classical low-flow, low-gradient (LFLG) AS with LVEF<50%, guidelines have recommend the use of low-dose dobutamine stress echocardiography to discriminate between true and pseudo-severe AS, with the recent ESC/EACTS guidance recommending DSE and/or aortic valve calcium score (AVCS) to aid diagnosis.^3–6^ Over the last decade, the AVCS measured by non-contrast cardiac computed tomography (CCT) has emerged as an adjunctive method to ascertain AS severity, with thresholds for AVCS >1300 in women and >2000 in men shown to have the best diagnostic accuracy for severe AS when compared to echocardiographic MG.^9,10^ Both ESC and AHA guidelines recommend the use of AVCS with these thresholds in patients with discordant AS associated with low flow. ^5,6^ However, these values were obtained in patients with a wide range of AS severities and may not reflect the threshold cut-off values in those being referred for TAVI.

CCT is also the current pre-procedural imaging modality of choice for pre-procedural planning of TAVI procedures, using multiphase cardiac CT angiography (CCTA) for evaluation of the aortic root and annulus. ^5.11^ This further offers the opportunity to evaluate the aortic valve area (CT-AVA) by direct planimetry in the mid-systolic phase of the cardiac cycle.^8,12^ The CT-AVA is an anatomical area similar to that obtained with transesophageal echocardiography and cardiovascular magnetic resonance imaging and has been shown to be larger than the EVA.^13^ Thus, much controversy exists about the optimal thresholds for severe AS both by AVA derived from direct planimetry (as CT-AVA) as well as EVA from the continuity equation.^4,7^

In this study, we aimed to evaluate the (i) optimal thresholds of AVCS and CT-AVA in patients with AS referred for TAVI assessment against the MG on TTE, (ii) the diagnostic performance of the derived AVCS thresholds compared to that of existing thresholds. We also evaluated the thresholds for EVA against the MG.

## METHODS

The study was a retrospective evaluation of CCT and TTE parameters of consecutive patients at three National Health Service (NHS) hospitals in England, United Kingdom, referred for TAVI assessment between 2010 to 2022. The study was approved by the Health Research Authority and Health and Care Research Wales (Reference: 21/HRA/1398).

Patients were excluded if there was a history of previous AVR or TAVI, moderate to severe aortic regurgitation, severe mitral regurgitation, or unavailable information on AVCS or MG. For CT-AVA evaluation, patients were excluded if image quality was suboptimal (Figure 1).

**Figure 1:**
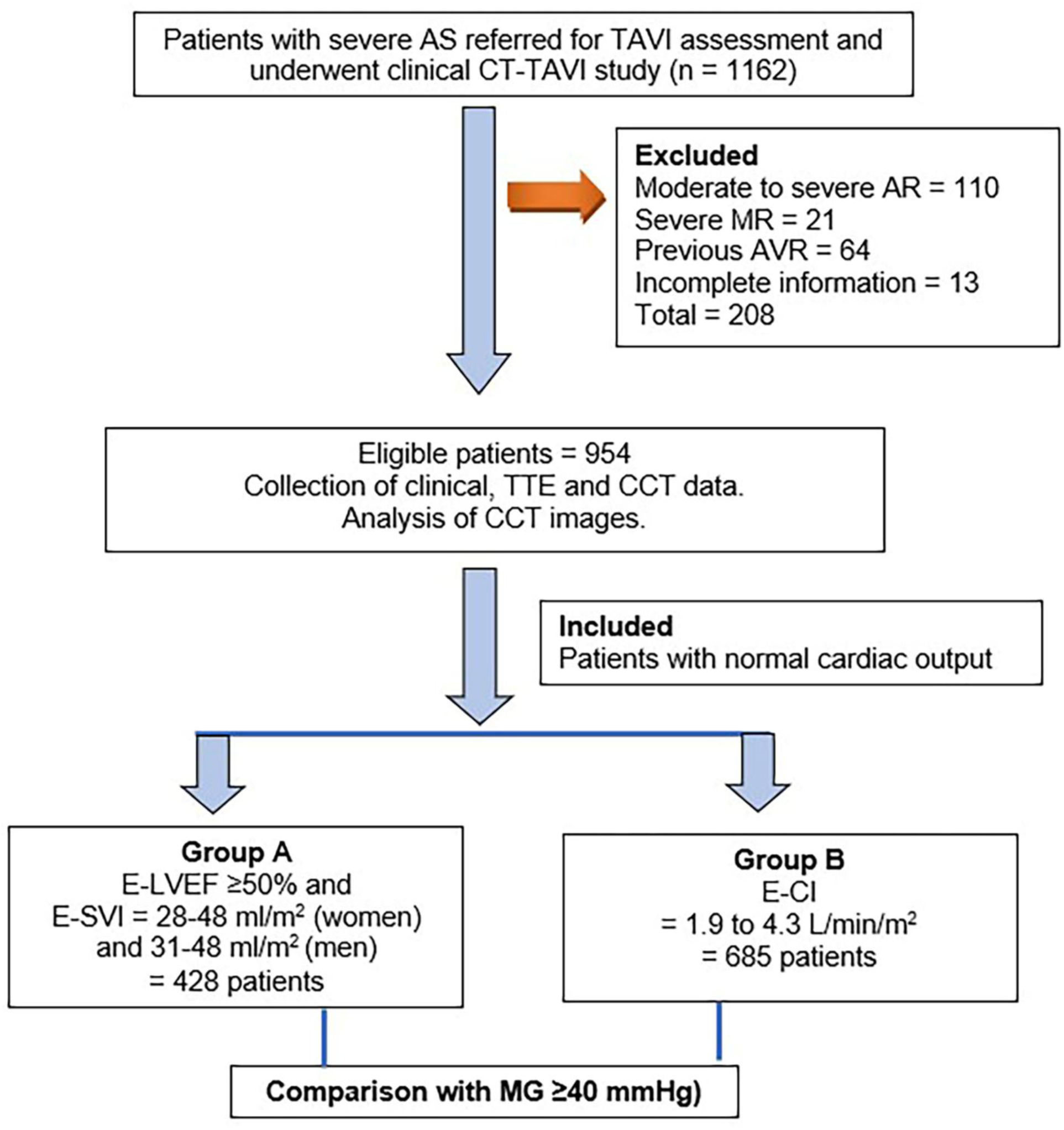
Flow graph demonstrating the recruitment of patient cohort. The figure demonstrates the number of patients excluded and the inclusion of patients in the two study groups. AR, aortic regurgitation; AS, aortic stenosis; CCT, cardiac computed tomography; E-CI, echocardiographic cardiac index; E-LVEF, echocardiographic left ventricular ejection fraction; MG, echocardiographic aortic valve mean gradient; E-SVI, echocardiographic stroke volume indexed; MR, mitral regurgitation; TTE, transthoracic echocardiography.

The remaining patients meeting two different criteria for normal stroke volume were considered for analysis in this study:

**Group A:** Echocardiographic left ventricular ejection fraction (E-LVEF) ≥50% and stroke volume index (E-SVI) = 28-48 ml/m^2^ in women and 31-48 ml/m^2^ in men,^14^ and

**Group B:** Normal cardiac index 1.9 to 4.3 L/min/m^2^.^15^

### CCT (Cardiac Computed Tomography)

The CCT scans were performed using similar clinical protocols on four different scanners. The CT scanners included Toshiba 64-detector (Toshiba Medical Systems Corporation, Japan), GE 128-slice (GE Healthcare, Chicago, USA), Siemens 128 slice (Siemens Healthineers, Erlangen, Germany), and Canon 320-detector (Canon Medical Systems Corporation, Otawara, Japan) at the three sites.

All CCT scans included an ECG-gated, non-contrast CT-AVCS and a contrast-enhanced CCTA scan. The CT-AVCS was performed using a standard protocol with 120 kVp, 3 mm contiguous acquisition, and prospective ECG-gating on all scanners. CCTA scans were performed using 100 or 120 kVp and 400-600 mAs depending upon the patient’s body size either with retrospective ECG-gating with dose-modulation or prospective ECG-gating between 0%-50% phases. Heart rate lowering drugs were not used due to a diagnosis of severe AS. 70-90 ml of non-ionic, low-osmolar contrast was administered intravenously.

Images were reconstructed at contiguous 1-2 mm collimation at 5-10% intervals throughout the cardiac cycle. The mean radiation dose was 10.1, 7.2 and 4.8 mSv (using a conversion factor of 0.014) for CT scans performed on 64-, 128-, and 320-slice CT scanners, respectively.

### CCT analysis

AVCS was quantified using the Agatston method restricted to the calcification within the AV leaflets with careful exclusion of calcium extending to the LVOT and the wall of the aortic root.^9^ AVCS-d (AVCS-density) was derived by dividing the AVCS by the annulus area obtained by direct planimetry. For the calculation of CT-AVA, CCTA data in the systolic phase (10-30%) were used. The method used to obtain and quantify the CT-AVA by direct planimetry in its true short-axis plane in mid-systole has been described previously.^8,12^ End-diastolic and end-systolic phases were used to obtain left ventricular volumes, stroke volume index (CT-SVI), and ejection fraction (CT-LVEF). The post-processing was performed by a level 2 or 3 SCCT cardiac radiologist or cardiologist, with >10 years’ experience using advanced post-processing software (TeraRecon, Durham, USA; Syngo Via, Siemens Healthineers, Erlangen, Germany; or Philips Medical Systems, Best, Netherlands). One of the senior authors (TKM) checked the AVCS and CT-AVA in 10-20% of cases from the other two institutions to ensure consistent methodology to minimise the inter-observer variability. All CT evaluations were performed without the knowledge of the TTE findings.

### Echocardiography

All patients underwent comprehensive TTE using commercially available echocardiography machines (Vivid-7 or 9, GE Healthcare, Chicago, USA; or IE33 or EPIQ CVx, Philips Healthcare, Andover, MA) using recommended methods for evaluation and diagnosis of AS.^3^ Left ventricular (LV) volumes and E-LVEF were measured and calculated using Simpson’s biplane method. LVOT diameter was measured from the parasternal long axis view in mid-systole near the annulus. LVOT velocity time integral (VTI) and mean velocity were measured using pulse wave Doppler in the apical 5-chamber view. The AV Vmax and MG were measured using continuous wave Doppler using multiple views to obtain the representative trace. The EVA was calculated using the continuity equation. Stroke volume was obtained by multiplying LVOT area by LVOT VTI and was indexed to body surface area (E-SVI).

In most patients, TTE was the first investigation resulting in the TAVI referral and TAVI-CT was performed within 6-months of TTE. In cases where a prior TTE was not available, the next earliest available pre-TAVI TTE was used for comparison.

### Statistical analysis

All analyses were performed separately for the two inclusion criteria for normal cardiac output (Groups A and B). Continuous variables are presented as mean ± SD or median (interquartile range) while categorical variables are presented as frequencies with percentages. The analyses were performed separately for males and females.

The predictive ability of AVCS, AVCS-d, CT-AVA, and EVA to determine severe AS (against MG ≥40 mmHg) was evaluated using ROC curves. The overall predictive ability was determined by the area under the ROC curve (AUC). ROC curves were also used to determine an optimal cut-point (threshold) for each parameter. The optimal cut-point was chosen as the point which maximised the combination of sensitivity and specificity (Youden index).

Logistic regression was performed to examine the predictive ability of both CT-AVCS and CT-AVA upon severe AS. The predicted regression model was used to create a single ‘combined’ variable, and a ROC curve analysis was performed on this combined variable as well. The area under the ROC curve of the combined variable was compared to each individual parameter using the methods suggested by DeLong, DeLong, and Clarke-Pearson.^16^ Further, the diagnostic accuracy of AVCS and AVCS-d at different thresholds was assessed against MG ≥40 mmHg.

All analyses were performed using STATA version 13.0 (StataCorp, College Station, Texas). A two-tailed p-value of < 0.05 was considered statistically significant

## RESULTS

Out of 1162 patients referred for TAVI CCT scans, 208 were excluded (Figure 1). Of the remaining 954 patients, inclusion criteria A and B were met by 428 patients and 685 patients, respectively. Mean age in both groups was 81± 7.5 years with women comprising 54% in Group A and 46% in Group B. Severe AS based on MG ≥40 mmHg was present in 239 (55.8%) of group A patients and 372 (54.4%) of group B patients. The prevalence of severe AS and other baseline characteristics is given in Table 1.

**Table 1:** Baseline characteristics of included patients.

|  | Inclusion criterion A (n = 428) |  |  | Inclusion criterion B (n = 685) |  |  |
| --- | --- | --- | --- | --- | --- | --- |
|  | All | Men | Women | All | Men | Women |
| Age, mean (±SD) | 81 (7.5) | 81 (7.7) | 81 (7.4) | 81 (7.5) | 81 (7.7) | 81 (7.2) |
| Sex (%) | 428 | 199 (47) | 229 (54) | 685 | 370 (54) | 315 (46) |
| BMI, mean (±SD) | 28 (6.1) | 28 (5.1) | 28 (6.8) | 27.9 (5.9) | 28 (6.5) | 28 (5.9) |
| Symptomatic (%) | 327 (76) | 156 (78) | 171 (75) | 542 (79) | 298 (81) | 244 (77) |
| Hypertension (%) | 249 (58) | 114 (57) | 135 (59) | 407 (60) | 214 (58) | 193 (61) |
| Diabetes mellitus (%) | 161 (38) | 79 (40) | 82 (36) | 270 (39) | 147 (40) | 123 (39) |
| Dyslipidaemia (%) | 166 (39) | 68 (34) | 98 (42) | 273 (40) | 136 (37) | 137 (43) |
| Smoking (%) | 122 (29) | 74 (38) | 48 (21) | 204 (30) | 129 (35) | 75 (24) |
| Known CAD (%) | 165 (39) | 93 (47) | 72 (31) | 274 (40) | 175 (47) | 99 (31) |
| Known AF (%) | 115 (27) | 67 (34) | 48 (21) | 183 (27) | 123 (33) | 60 (19) |
| Known heart failure (%) | 57 (13) | 32 (16) | 25 (11) | 131 (19) | 88 (24) | 43 (14) |
| AVCS AU, median (IQR) | 2424 (1686, 3609) | 3055 (2168, 4364) | 2047 (1380, 3032) | 2519 (1700-3900) | 2983 (2092, 4314) | 2141 (1406, 3233) |
| Tricuspid AV (%) | 357 (86) | 158 (82) | 199 (89) | 555 (83) | 291 (81) | 264 (86) |
| CT-AVA, cm<sup>2</sup> (mean ± SD) | 0.97 (0.3) | 1.0 (0.3) | 0.9 (0.3) | 0.90 (0.3) | 1.0 (0.3) | 0.9 (0.3) |
| CT-AVA ≤1.0 cm<sup>2</sup> (%) | 253 (59) | 96 (48) | 157 (69) | 398 (58) | 183 (49) | 215 (68) |
| E-Vmax, m/s (mean ± SD) | 4.3 (0.7) | 4.1 (0.7) | 4.4 (0.8) | 4.2 (0.8) | 4.1 (0.7) | 4.3 (0.8) |
| E-Vmax ≥4 m/s (%) | 286 (67) | 123 (62) | 163 (71) | 445 (65) | 225 (61) | 220 (70) |
| E-MG, mmHg (mean ± SD) | 44.1 (16) | 42 (13) | 46 (18) | 41 (17) | 39 (15) | 43 (18) |
| E-MG ≥40 mmHg (%) | 239 (56) | 104 (52) | 135 (59) | 372 (54) | 183 (49) | 189 (60) |
| EVA, cm<sup>2</sup> (mean ± SD) | 0.72 (0.18) | 0.76 (0.18) | 0.67 (0.18) | 0.71 (0.2) | 0.76 (0.2) | 0.7 (0.2) |
| EVA ≤1.0 cm<sup>2</sup> (%) | 405 (95) | 186 (94) | 219 (96) | 637 (93) | 341 (92) | 296 (94) |
| DI (mean ± SD) | 0.23 (0.06) | 0.23 (0.06) | 0.24 (0.07) | 0.22 (0.07) | 0.22 (0.08) | 0.23 (0.07) |
| DI ≤0.25 (%) | 275 (64) | 135 (68) | 140 (61) | 446 (65) | 253 (68) | 193 (61) |
| E-LVEF, % (mean ± SD) | 62.9 (7.8) | 61.4 (7.5) | 64.2 (7.8) | 60 (13) | 58 (14) | 61 (12) |
| E-SVI, (mean ± SD) | 38.7 (5.1) | 39 (4.9) | 38 (5.3) | 38.7 (8.9) | 38.5 (8.9) | 38.7 (9.1) |
| Cardiac index, L/min/m<sup>2</sup> | 2.8 (0.7) | 2.7 (0.8) | 2.9 (0.61) | 2.8 (0.6) | 2.7 (0.6) | 2.9 (0.6) |

Shaded rows represent the prevalence of severe AS.
AF, atrial fibrillation; AV, aortic valve; AVCS, aortic valve calcium score; AU, Agatston units; BMI, Body mass index; CAD, coronary artery disease; CT-AVA, CT aortic valve area; DI, dimensionless index; E-LVF, echocardiographic left ventricular ejection fraction, E-MG, echocardiographic aortic valve mean gradient; E-SVI, echocardiographic stroke volume indexed; EVA, effective valve area; E-Vmax, echocardiographic aortic valve velocity maximum

In Group A, the optimal thresholds of AVCS for predicting a MG ≥40 mmHg were ≥3046 AU in men and ≥2034 AU in women. For AVCS-d, the corresponding thresholds were ≥ 712 AU in men and ≥ 443 AU in women (Table 2). In Group B, the best threshold for AVCS were ≥2983 AU in men and ≥1962 AU in women and for AVCS-d ≥ 639 AU in men and ≥ 490 AU in women (Table 3).

**Table 2:**
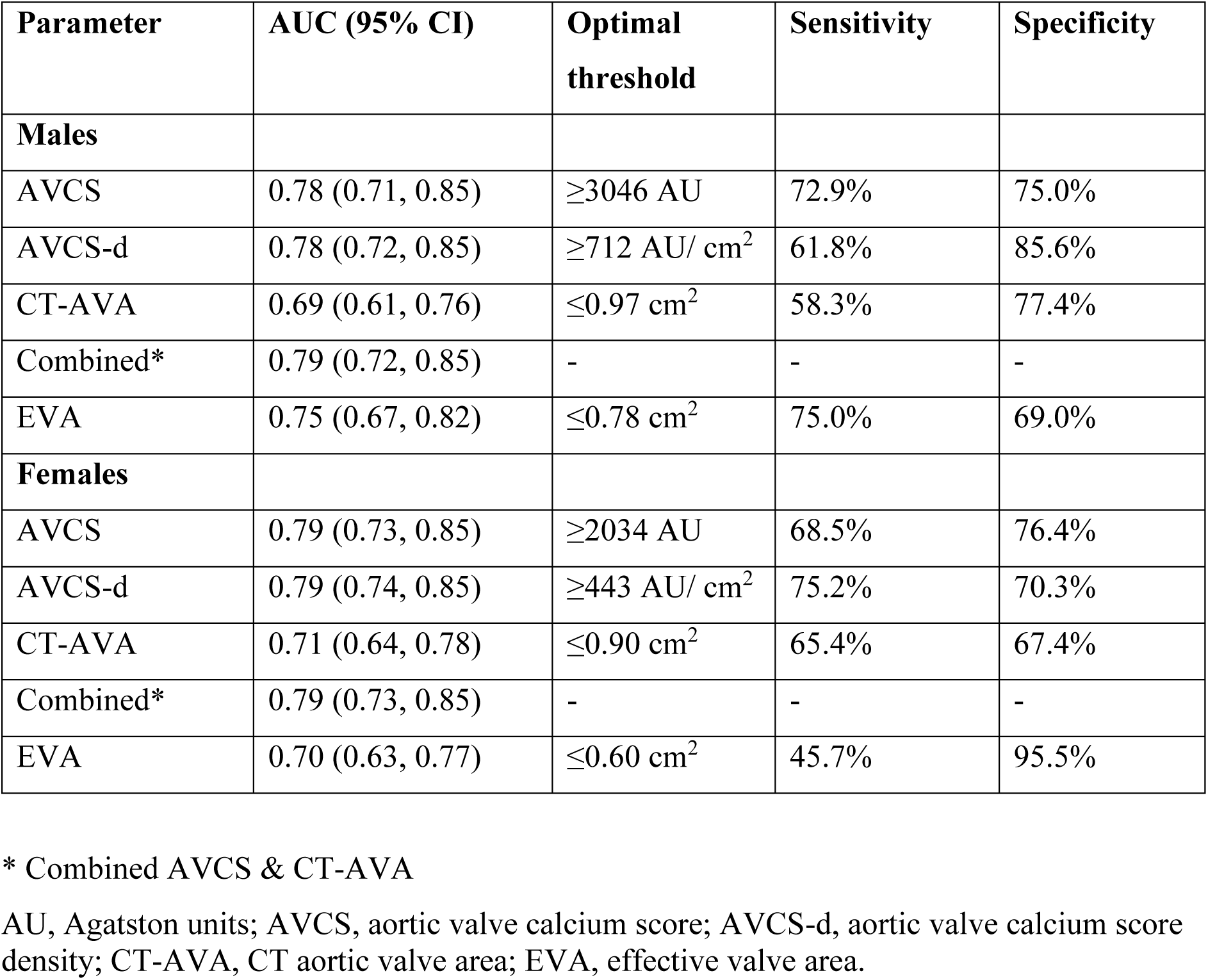
Group A: Optimal thresholds for the prediction of severe AS (MG≥40 mmHg)

**Table 3:**
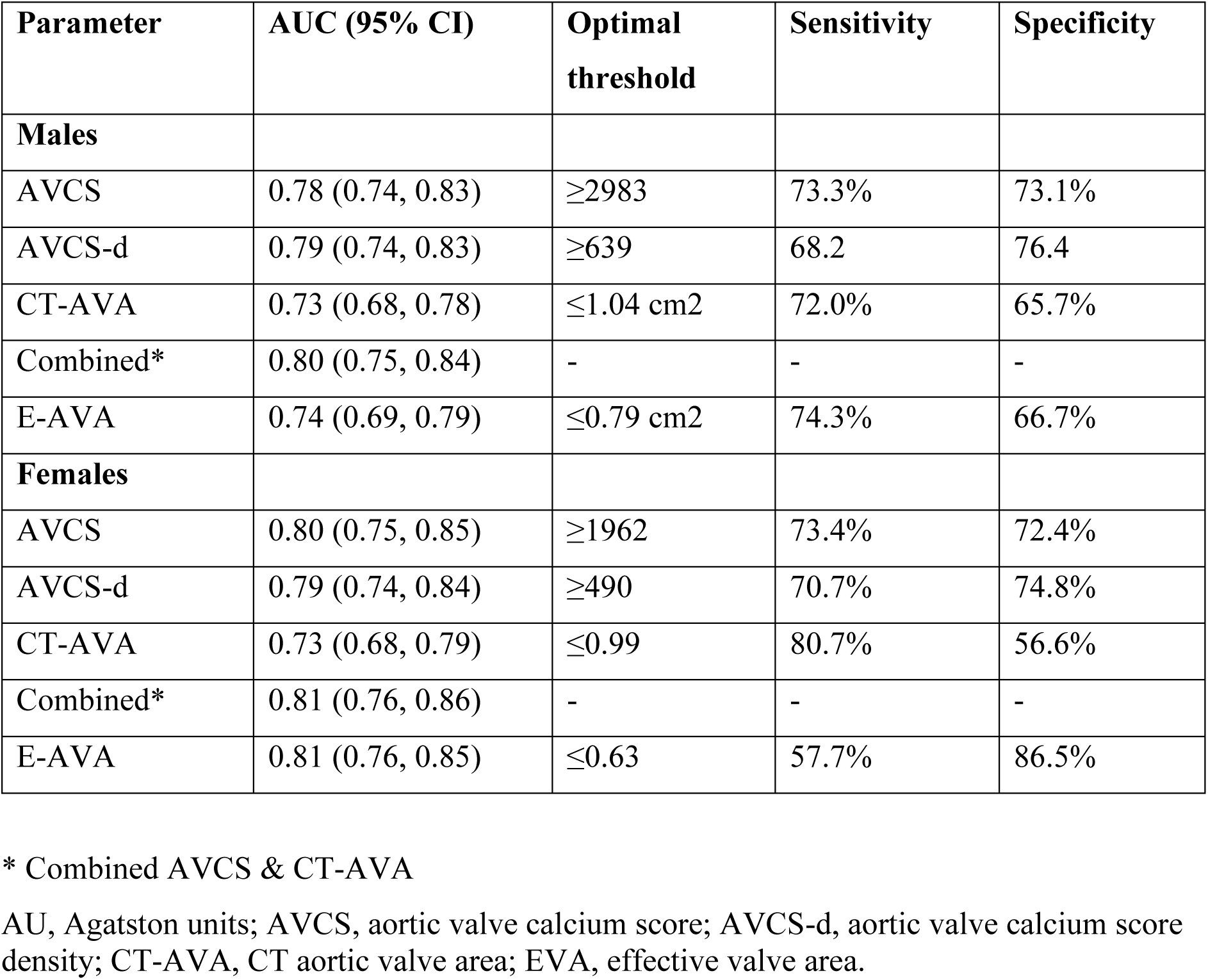
Group B: Optimal cut-points (thresholds) for the prediction of severe AS (MG≥40 mmHg)

The best threshold for CT-AVA was ≤0.97 cm^2^ in men in group A (AUC 0.69, 95% confidence interval 0.61-0.76) and ≤1.04 cm^2^ in group B (AUC 0.73, 95% confidence interval 0.68-0.78). In women, the threshold was ≤0.90 cm^2^ in Group A and ≤0.99 cm^2^ in Group B.

For EVA, the best threshold in men was ≤0.78 cm^2^ in Group A (AUC 0.78, 95% confidence interval 0.67-0.82) and ≤0.79 cm^2^ in Group B (AUC 0.74, 95% confidence interval 0.69-0.79). In women, the thresholds were ≤0.60 cm^2^ in Group A and ≤0.63 cm^2^ in Group B.

As a single parameter, AVCS had the best predictive value at the above threshold values with an AUC in men of 0.78 (95% confidence interval, 0.71-0.85) in both groups, while in women it had an AUC of 0.79 (95% confidence interval, 0.73-0.85) and 0.80 (95% confidence interval, 0.75-0.85) in Groups A and B, respectively. The AUC for AVCS and AVCS-d did not vary between the two inclusion criteria. A combination of AVCS and CT-AVA had similar predictive value to AVCS alone in Group A while the combination improved the AUC in Group B from 0.78 to 80 in men and from 0.80 to 0.81 in women.

Table 4 demonstrates the prevalence of severe AS based on the absolute AVCS and AVCS-d as per the best thresholds obtained in this study and their diagnostic accuracy obtained in this study and compares the prevalence and diagnostic accuracy based on the thresholds reported in previous studies. The thresholds obtained in this study are much higher providing a greater specificity and positive predictive value resulting in a higher AUC.

**Table 4:** Diagnostic accuracy of derived thresholds for AVCS, CT-AVA, and EVA compared to existing thresholds for the prediction of severe AS based on MG ≥40 mmHg in Group B.

| Parameter | Prevalence | AUC | Sensitivity | Specificity | PPV | NPV |
| --- | --- | --- | --- | --- | --- | --- |
| **AVCS** |  |  |  |  |  |  |
| **Men** |  |  |  |  |  |  |
| AVCS $\geq 3000^*$ | 48% | 0.78 | 72% | 74% | 73% | 74% |
| AVCS $\geq 2062$ | 74% | 0.61 | 93% | 42% | 61% | 86% |
| AVCS-d $\geq 639^*$ | 45% | 0.79 | 68% | 76% | 74% | 71% |
| AVCS-d $\geq 500$ | 61% | 0.61 | 83% | 58% | 66% | 77% |
| **Women** |  |  |  |  |  |  |
| AVCS $\geq 2000^*$ | 53% | 0.80 | 72% | 72% | 80% | 63% |
| AVCS $\geq 1300$ | 78% | 0.64 | 94% | 39% | 70% | 80% |
| AVCS-d $\geq 490^*$ | 51% | 0.79 | 71% | 75% | 81% | 63% |
| AVCS-d $\geq 420$ | 64% | 0.65 | 82% | 59% | 75% | 69% |
| **CT-AVA $\leq 1 \text{ cm}^2^*$** | 59% | 0.69 | 77% | 63% | 73% | 68% |
| **EVA** |  |  |  |  |  |  |
| EVA $\leq 1 \text{ cm}^2$ | 95% | 0.55 | 99% | 11% | 59% | 96% |
| EVA $\leq 0.8 \text{ cm}^2$<br>(men) * | 50% | 0.67 | 80% | 54% | 63% | 74% |
| EVA $\leq 0.6 \text{ cm}^2$<br>(women) * | 60% | 0.67 | 56% | 85% | 85% | 56% |

\* Thresholds for current study
AVCS, aortic valve calcium score; AVCS-d, aortic valve calcium score density; CT-AVA, CT aortic valve area; EVA, effective valve area.

## DISCUSSION

This study provides the optimal AVCS thresholds for the diagnosis of severe AS in patients clinically referred for TAVI, using two different criteria to define normal cardiac output. The optimal thresholds are approximately 3000 AU in men and 2000 AU in women and provide the best predictive value for severe AS using MG ≥40 mmHg as the reference standard. This study also identifies the optimal thresholds for CT-AVA for the diagnosis of severe AS as being ≤1.0 cm^2^ in males and ≤0.9 cm^2^ in females, and for EVA being ≤0.8 cm^2^ in men and ≤0.6 cm^2^ in women.

Degenerative disease is the most common aetiology of AS with a complex pathobiology involving multiple features such as fibrosis, inflammation, oxidative stress, angiogenesis, haemorrhage, and osteogenic differentiation leading ultimately to calcification.^17^ Two early studies evaluating AVCS and AS severity reported high thresholds of 3700 AU when compared to peak velocity in 157 patients.^18^ and 2500 AU compared to catheter peak to peak and mean gradients in 72 patients.^19^

The first large study to report AVCS threshold included 451 patients with SVI >35 ml/m^2^ in patients with moderate to severe AS with concordant EVA and MG grading and prevalence of severe AS by MG ≥40 mmHg of 44%.^9^ This study was the first to demonstrate a sex difference in the AVCS load related to AS severity with the best thresholds in men and women being ≥2065 AU (AUC, 0.90) and ≥1275 AU (AUC, 0.91), respectively. The second large study included 437 out of 918 patients with concordant severe grading (EVA ≤1cm^2^ and Vmax ≥4m/s) and found best AVCS thresholds as ≥2062 AU (AUC, 0.89) and ≥1377 AU (AUV, 0.92), respectively.^10^ When compared to the best AVCS threshold of ≥3000 AU in men obtained in this study, an AVCS threshold of ≥2062 reduces the AUC from 0.74 to 0.61 due to reduction of specificity from 73% to 45% (Table 4). Similarly, there is a reduction in diagnostic accuracy in women as well, indicating that the originally described optimal AVCS thresholds perform less well in patients referred for TAVI. The reasons for much higher AVCS thresholds in our cohort are uncertain but could possibly be due to higher age in our patient cohort referred for TAVI (81±7.5 years) compared to other studies (75±12 years) and different contributions of calcification and fibrosis to the severity of AS in the study cohorts. It may also conceivably relate to differences in calcification levels in patients with symptomatic versus asymptomatic severe AS.

Measuring AVA is considered ideal for assessing severe AS due to it being the least flow dependent parameter compared to Vmax and MG.^20^ The AVA was initially derived using Gorlin’s formula by measuring the transvalvular aortic pressure gradient by cardiac catheterisation but was superseded in routine practice by use of the continuity equation-derived EVA on TTE, as the latter is non-invasive.^21^ However, both are indirect measurements and affected by flow across the left ventricular outflow tract.^22,23^ The fact that EVA is generally smaller compared to AVA derived from planimetry techniques has been highlighted by a number of publications and can cause discordance in AS severity in up to 30% of patients.^4,9,23^ In this study, EVA ≤1cm^2^ over-diagnosed severe AS in 40% patients when compared to MG ≥40 mmHg. It is now well known that the aortic annulus or LVOT is mostly elliptical in shape, and it is the measurement of a smaller diameter on TTE that is the most likely cause of underestimation of EVA.^24,25^ The discrepancy between EVA and other TTE parameters was highlighted in a study of 2427 patients with a diagnosis of severe AS based upon EVA <1cm^2^, Vmax >4 m/s or MG >40 mmHg.^26^ By calculating curve fits for the relationship between EVA and MG using the Gorlin equation and between EVA and Vmax using the continuity equation, authors found that an EVA of 1 cm^2^ corresponds to a MG of 21 mmHg and Vmax of 3.3 m/s. Conversely, a MG of 40 mmHg and Vmax of 4 m/s corresponded to an EVA of 0.75 cm^2^ and 0.82 cm^2^. In the present study, the MG of 40 mmHg correlates best with EVA of 0.8 cm^2^ in men and 0.6 cm^2^ in women, which matches well with the thresholds proposed by Minners et al for EVA. ^26^

This study also shows that EVA ≤1cm^2^ is a highly sensitive parameter for severe AS but has low specificity. It is best to confirm the AS severity by Vmax/MG after normalising the stroke volume or with CT parameters. However, the ESC and AHA guidelines continue to use the EVA ≤1 cm^2^ criterion due to its prognostic value in many studies.^5,6^

CT-AVA can be routinely obtained on CCTA images if ECG-gating includes the mid-systolic phases of the cardiac cycle (typically 10-30%). CCTA images are volumetric with a high spatial resolution (0.3-0.6 mm) and can be easily aligned to the best short-axis plane of the AV valve on the CT software after the scan is acquired.^8,12,13,27^ One can thus obtain an accurate anatomical AVA that is the least flow dependent AS parameter provided the correct method is followed.^8,12^ AV calcification does not hamper planimetry as it tends to occur along the inner edges of the valve cusps. However, suboptimal images can be obtained if there is inadequate contrast density or in the presence of atrial fibrillation. If the CCTA scan covers the whole cardiac cycle, one can also quantify the stroke volume and LVEF from the same scan.

Several authors have attempted to combine the haemodynamic TTE parameters with LVOT/annulus area from CCTA to overcome the limitation of using a single, smaller LVOT diameter measured on TTE.^28,29^ or use a correction factor.^30^ However, in this study we have focused on the CT-AVA by direct planimetry as it represents a direct anatomical method.

### Strengths and Limitations

There are several strengths to our study. Firstly, it comprises of a large number of patients with diagnosis of severe AS on TTE for consideration of TAVI procedure where CCT is now routinely performed. Secondly, it is a multi-centre study with both TTE and CCT performed on different scanners with CCT analysed by an experienced investigator at each site. Thirdly, the study has only considered patients with normal stroke volume index or normal cardiac index for estimation of best thresholds, as the latter may be affected with low flow states.

Finally, along with AVCS, the study has also assessed the best thresholds for CT-AVA and EVA in patients with normal flow.

The limitation of the study includes its retrospective design with comparison of TTE and CCT scans clinically performed within 6-months. All attempts were made to exclude patients where there was signficant clinical deterioration between the TTE and CCT scans.

Nevertheless, even if this was the case, the CCT would have likely shown an increased prevalence of severe AS as it was performed after the TTE that had formed the basis of TAVI assessment referral.

Like any parameter, the accuracy of AVCS and CT-AVA depends upon the experience of the interpreting physician and use of correct methodology. In a previous study, two of the authors with high experience have tested inter-observer reproducibility.^8^ One of the same authors has rechecked the AVCS and CT-AVA in up to 20% of patients as stated in the Methods section.

We have taken MG as the “gold-standard” in this study comprising of patients with normal flow and cardiac output. However, it is recognised that MG can be subject to various errors resulting in both under and over-estimation.^7^

## Conclusion

The optimal CT-AVCS thresholds for severe AS in patients referred for TAVI are much higher compared to previously reported thresholds, suggesting a need for re-evaluation of the criteria used in this patient population. This study also establishes the optimal cut-off for CT-AVA and EVA in patients with normal stroke volume and cardiac output.

## Data Availability

The study data will be available to any investigator on appropriate request.

## Acknowledgements

The authors acknowledge the support provided by Paul Bassett for the statistical analysis.

## Nonstandard Abbreviations and Acronyms

AS: Aortic stenosis
AVA: Aortic valve area
AVCS: Aortic valve calcium score
CCT: Cardiac Computed Tomography
CCTA: Cardiac Computed Tomography Angiography
EVA: Effective valve area
MG: Mean gradient Vmax, Peak velocity
TAVI: Transcatheter Aortic Valve Implantation
TTE: Transthoracic Echocardiography

